# Epidemiological characteristics and transmission dynamics of the outbreak caused by the SARS-CoV-2 Omicron variant in Shanghai, China: a descriptive study

**DOI:** 10.1101/2022.06.11.22276273

**Authors:** Zhiyuan Chen, Xiaowei Deng, Liqun Fang, Kaiyuan Sun, Yanpeng Wu, Tianle Che, Junyi Zou, Jun Cai, Hengcong Liu, Yan Wang, Tao Wang, Yuyang Tian, Nan Zheng, Xuemei Yan, Ruijia Sun, Xiangyanyu Xu, Xiaoyu Zhou, Shijia Ge, Yuxiang Liang, Lan Yi, Juan Yang, Juanjuan Zhang, Marco Ajelli, Hongjie Yu

**Author notes:** Co-corresponding authors: Juanjuan Zhang, School of Public Health, Fudan University, Key Laboratory of Public Health Safety, Ministry of Education, Shanghai 200032, China.; Hongjie Yu, School of Public Health, Fudan University, Key Laboratory of Public Health Safety, Ministry of Education, Shanghai 200032, China. These authors contributed equally to this work (Zhiyuan Chen, Xiaowei Deng, Liqun Fang).

## Abstract

**Background:** In early March 2022, a major outbreak of the severe acute respiratory syndrome coronavirus 2 (SARS-CoV-2) Omicron variant spread rapidly throughout Shanghai, China. Here we aimed to provide a description of the epidemiological characteristics and spatiotemporal transmission dynamics of the Omicron outbreak under the population-based screening and lockdown policies implemented in Shanghai.

**Methods:** We extracted individual information on SARS-CoV-2 infections reported between January 1 and May 31, 2022, and on the timeline of the adopted non-pharmacological interventions. The epidemic was divided into three phases: i) sporadic infections (January 1–February 28), ii) local transmission (March 1–March 31), and iii) city-wide lockdown (April 1 to May 31). We described the epidemic spread during these three phases and the subdistrict-level spatiotemporal distribution of the infections. To evaluate the impact on the transmission of SARS-CoV-2 of the adopted targeted interventions in Phase 2 and city-wide lockdown in Phase 3, we estimated the dynamics of the net reproduction number (*R*_*t*_).

**Findings:** A surge in imported infections in Phase 1 triggered cryptic local transmission of the Omicron variant in early March, resulting in the largest coronavirus disease 2019 (COVID-19) outbreak in mainland China since the original wave. A total of 626,000 SARS-CoV-2 infections were reported in 99.5% (215/216) of the subdistricts of Shanghai. The spatial distribution of the infections was highly heterogeneous, with 40% of the subdistricts accounting for 80% of all infections. A clear trend from the city center towards adjacent suburban and rural areas was observed, with a progressive slowdown of the epidemic spread (from 544 to 325 meters/day) prior to the citywide lockdown. During Phase 2, *R*_*t*_ remained well above 1 despite the implementation of multiple targeted interventions. The citywide lockdown imposed on April 1 led to a marked decrease in transmission, bringing *R*_*t*_ below the epidemic threshold in the entire city on April 14 and ultimately leading to containment of the outbreak.

**Interpretation:** Our results highlight the risk of widespread outbreaks in mainland China, particularly under the heightened pressure of imported infections. The targeted interventions adopted in March 2022 were not capable of halting transmission, and the implementation of a strict, prolonged city-wide lockdown was needed to successfully contain the outbreak, highlighting the challenges for successfully containing Omicron outbreaks.

**Funding:** Key Program of the National Natural Science Foundation of China (82130093).

**Research in context:** *Evidence before this study:* On May 24, 2022, we searched PubMed and Europe PMC for papers published or posted on preprint servers after January 1, 2022, using the following query: (“SARS-CoV-2” OR “Omicron” OR “BA.2”) AND (“epidemiology” OR “epidemiological” OR “transmission dynamics”) AND (“Shanghai”). A total of 26 studies were identified; among them, two aimed to describe or project the spread of the 2022 Omicron outbreak in Shanghai. One preprint described the epidemiological and clinical characteristics of 376 pediatric SARS-CoV-2 infections in March 2022, and the other preprint projected the epidemic progress in Shanghai, without providing an analysis of field data. In sum, none of these studies provided a comprehensive description of the epidemiological characteristics and spatiotemporal transmission dynamics of the outbreak.

*Added value of this study:* We collected individual information on SARS-CoV-2 infection and the timeline of the public health response. Population-based screenings were repeatedly implemented during the outbreak, which allowed us to investigate the spatiotemporal spread of the Omicron BA.2 variant as well as the impact of the implemented interventions, all without enduring significant amounts of underreporting from surveillance systems, as experienced in other areas. This study provides the first comprehensive assessment of the Omicron outbreak in Shanghai, China.

*Implications of all the available evidence:* This descriptive study provides a comprehensive understanding of the epidemiological features and transmission dynamics of the Omicron outbreak in Shanghai, China. The empirical evidence from Shanghai, which was ultimately able to curtail the outbreak, provides invaluable information to policymakers on the impact of the containment strategies adopted by the Shanghai public health officials to prepare for potential outbreaks caused by Omicron or novel variants.

## Background

The first wave of coronavirus disease 2019 (COVID-19) in China subsided quickly with strict containment measures in March 2020.^1^ As numerous variants have emerged across the globe, China successfully contained multiple COVID-19 outbreaks by adhering to a containment policy. This policy aimed to curb flare-ups of local transmissions in the shortest possible time, which is done by relying on a set of non-pharmacological interventions (NPIs) with adjusted intensities according to the situation on the ground.^2,3^

In November 2021, the severe acute respiratory syndrome coronavirus 2 (SARS-CoV-2) Omicron variant was first reported in South Africa.^4^ Its high transmissibility and immune escape properties enabled it to rapidly replace previous strains and become dominant globally.^5,6^ Although the containment policy implemented in China had been effective in containing pre-Omicron outbreaks, its effectiveness against Omicron was unclear. In fact, a total of 750,000 SARS-CoV-2 infections were reported in mainland China in the first five months of 2022,^7^ the majority in Shanghai.

Shanghai is one of the most populous and economically advanced metropolises in China, with a population of nearly 25 million^8^ and unique features compared to other areas in mainland China and abroad. The population in Shanghai was mostly vaccinated with domestically-developed inactivated vaccines, which had relatively lower effectiveness in preventing SARS-CoV-2 infections due to their lower antibody-neutralizing responses compared to mRNA vaccines^9,10^. In addition, the vaccination coverage was highest in the young population (100.0% in individuals aged between 12 and 17 years had completed the primary vaccine schedule as of March 2022)^11^ and lowest in the elderly (only 62.0% in individuals aged 60 years or older had completed the primary schedule as of April 2022).^12^ Compared to the rest of the cities in mainland China, Shanghai had the highest risk of SARS-CoV-2 infection importation due to its global connectivity: about 30-40% of all international flights arriving in China since 2020 landed in Shanghai.^13^ Although a quarantine period of two weeks was mandatory for all incoming travelers, lapses in these measures may have led to the repeated seeding of SARS-CoV-2 into the community.^14,15^ Consequently, in early March 2022, a major outbreak of the Omicron BA.2 variant started to spread in Shanghai.^16^

In this study, we provide a detailed description of Shanghai’s outbreak response timeline, the spatiotemporal distribution of COVID-19 cases, and Omicron transmission dynamics since 2022. The analysis of the Shanghai 2022 Omicron outbreak could provide invaluable information to policymakers on the impact of the containment strategies adopted by Shanghai to prepare for potential outbreaks caused by Omicron or future novel variants.

## Methods

### Definition of infection and case

An asymptomatic infection is defined as a PCR-confirmed individual who i) does not meet any of the following clinical criteria: fever, cough, sore throat, and other self-perceived and clinical-identifiable symptoms or signs; and ii) has no radiographic evidence of pneumonia. Laboratory-confirmed cases are categorized into four types based on clinical severity: mild, moderate, severe, and critical cases, with detailed definitions in the **Supplementary Information**. Detailed surveillance, detection, and management of infections and cases are also presented in the **Supplementary Information**.

### Public health response

We summarize the NPIs implemented in Shanghai before and during the Omicron waves. Prior to the initial surge in Omicron, Shanghai maintained a baseline level of intensive NPIs against potential outbreaks, including stringent border control policies, symptom-based surveillance, case isolation, tracing of close contacts (requires quarantine in separate facilities) and contacts of contacts, occupation-based screening, targeted screening of individuals at high risk of infection (e.g., contacts of contacts), and a set of other social distancing measures such as travel restrictions and community confinement (**Figure 1**). After the Omicron variant was introduced in Shanghai, a set of additional NPIs were implemented to halt its transmission. In terms of border control measures, the point of entry for international flights to Shanghai was diverted to other airports. At the local scale, the closure of in-person educational activities at all levels was implemented from March 12 to 15, 2022 (starting from primary and secondary school, then followed by universities). From March 16 to March 27, Shanghai adopted grid management at the subdistrict level, fractioning subdistricts into high-risk areas. The lists of high-risk areas changed dynamically over time based on the following principles: the epidemiological situation measured in terms of number of cases and infections, gathering of crowds, population density, social characteristics, and economic activity.^17^ During this period, several rounds of nucleic acid screening were performed within high-risk and non-high-risk areas. In high-risk areas, either one or two rounds of population-based nucleic acid screening were performed within 48 hours, together with a lockdown. In non-high-risk areas, a single round of mass PCR screening was performed between March 18 and 20. Afterwards, the entire city entered a phased stage of lockdown, where eastern Shanghai (which comprises areas east of the Huangpu River, **Supplementary Figure 1**) entered a population-wide lockdown on March 28, and then the rest of Shanghai entered the lockdown phase on April 1. The lockdown restriction was lifted on June 1 when the daily number of infections first declined to 10..

**Figure 1.**
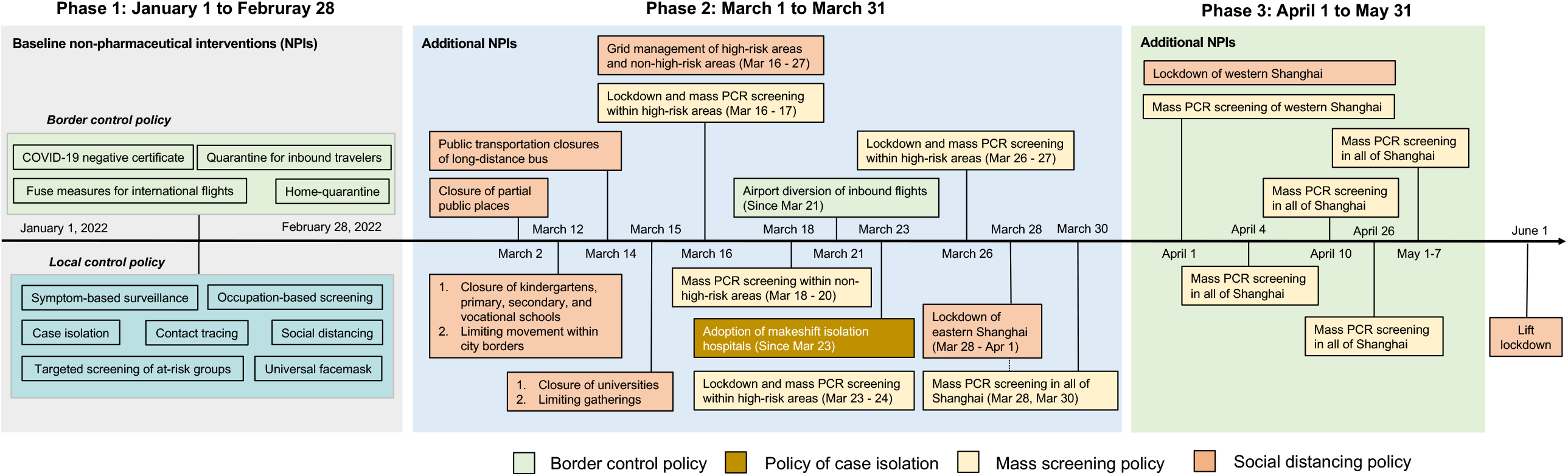
Timeline of the public health response in Shanghai by epidemic phase.

### Data sources and collection

Aggregated data on the number of local and imported infections according to symptomatic status in Shanghai from December 2019 were gathered from the Shanghai Municipal Health Commission through a combined approach of manual downloads and compiled scripts. Individual line lists of all SARS-CoV-2 infections were retrieved from multiple publicly available official data sources (websites of municipal health commissions) and integrated with the supplementary information gathered from the websites of local government media (**Supplementary Table 1**). The resulting line list contains the following variables: residential location (address, district, and subdistrict), symptomatic status (including the clinical outcomes of initially asymptomatic infections), date of official reporting, and means of infection identification (i.e., routine screening of community groups or general screening of the quarantined population) (**Supplementary Table 2**). Specifically, community screening groups referred to those residents living in areas reporting no new infections, people proactively looking for healthcare or nucleic acid tests, and essential workers. The quarantined population corresponded to individuals who were quarantined at home or in designated facilities, lived in areas reporting new infections or other key settings under close management, and were identified through screening of close contacts^18^. We obtained the timeline of the adopted NPI strategies from the official announcements of the authorities of Shanghai through search engine queries; the list of high-risk areas during March 16 and March 27 was found by searching on local media pages (**Supplementary Table 3**). More details on data collection are provided in the **Supplementary Information**.

### Statistical analyses

#### Definition of the three phases of the epidemic

Three phases of the epidemic were defined according to the staged NPIs policies adopted and the epidemic situation. The first phase covers the period from January 1 to February 28, 2022, during which the main risk was posed by the importation of infections and only small numbers of sporadic and locally transmitted cases were recorded. The second phase started on March 1, immediately before the outbreak of the BA.2 Omicron variant was confirmed^16^. In addition, no epidemiological link between infections reported before and after March 1, 2022, was identified by the authorities. The third phase covers the period from April 1 to May 31, 2022, when the entire city went into staged lockdowns and the epidemic started to subside.

#### Inference of delay between sampling and reporting

The date of symptom onset or infection was needed to describe the transmission dynamics, whereas we only had the date of official reporting for each infection. We estimated the delayed days between the sampling dates and reporting dates and then generated the sampling date for each infection by assuming that the sampling dates were close to the dates of symptom onset or infection time.

The mean reporting delay was determined by computing the cross-correlation coefficients between the cumulative number of reported SARS-CoV-2 infections and the cumulative number of sampled specimens under different lengths of lag (**Supplementary Figure 2**), the details of which are presented in the **Supplementary Information**. We applied delays of 2 days, 3 days, and 2 days for the periods before March 15, between March 16 and May 14, and after May 15, respectively. In the sensitivity analysis, we considered a four-day delay for the period from March 16 to May 14.

In addition to reporting delay, we also estimated individual locations and the progression of pre-symptomatic infections for data analysis (see **Supplementary Information** for details).

#### Spatial trends

Trend surface analysis was used to explore the SARS-CoV-2 spread in Shanghai, China. Thin-plate spline regression was used to interpolate the first date of sampling in each 3 km × 3 km cell of a gridded population of Shanghai to draw a spatial trend surface plot. The local slope (time/distance) of the trend surface was then measured using a 3 × 3 moving window filter, and the inverse of the slope value was used to estimate the speed of the spatial spread of SARS-CoV-2 (distance/time).^20,21^

#### Estimation of epidemiological parameters

The net reproduction number (*R*_*t*_) was estimated using the EpiEstim R package.^22^ Briefly, the estimate is based on a Bayesian approach that relies on the knowledge of the time series of infections (sampling date of positive tests in our analysis) and the distribution of the generation time. In the absence of estimates of the generation time for the analyzed outbreak, we assumed it to be gamma-distributed with a mean of 2.72 days (shape = 3.25, scale = 0.84), corresponding to the estimated serial interval during the Omicron wave in Hong Kong in the presence of strict interventions.^19^

We estimated the growth rate (r) and 95% confidence interval (CI) in the second epidemic phase by fitting a linear regression to the logarithm of the incidence of infections. The doubling time in the early stage was then estimated as ln(2)/r, where the 95% CI was calculated using the delta method. All analyses and visualization were performed in ArcGIS 10.7 and R (version 4.0.2).

#### Role of the funding source

The funder of the study had no role in study design, data collection, data analysis, data interpretation, or writing of the report. The corresponding authors had full access to all the data in the study and had final responsibility for the decision to submit for publication.

This study was approved by the institutional review board of the School of Public Health, Fudan University (IRB# 2022-05-0968). All data were collected from publicly available sources. Data were de-identified, and the need for informed consent was waived.

## Results

Following the successful containment of the initial epidemic wave caused by the Wuhan outbreak in early 2020, Shanghai was under constant pressure of potential outbreak introductions due to a high influx of international travelers relative to the rest of the cities in mainland China. Despite ranking first in the cumulative number of imported confirmed SARS-CoV-2 infections before 2022, Shanghai had sustained minimum local transmission prior to the emergence of the Omicron variant (**Figure 2a**), which was attributable to well-managed entry quarantine, screenings, and case isolation programs. Nonetheless, sporadic local transmissions events occurred, and all pre-Omicron outbreaks were swiftly suppressed.

**Figure 2.**
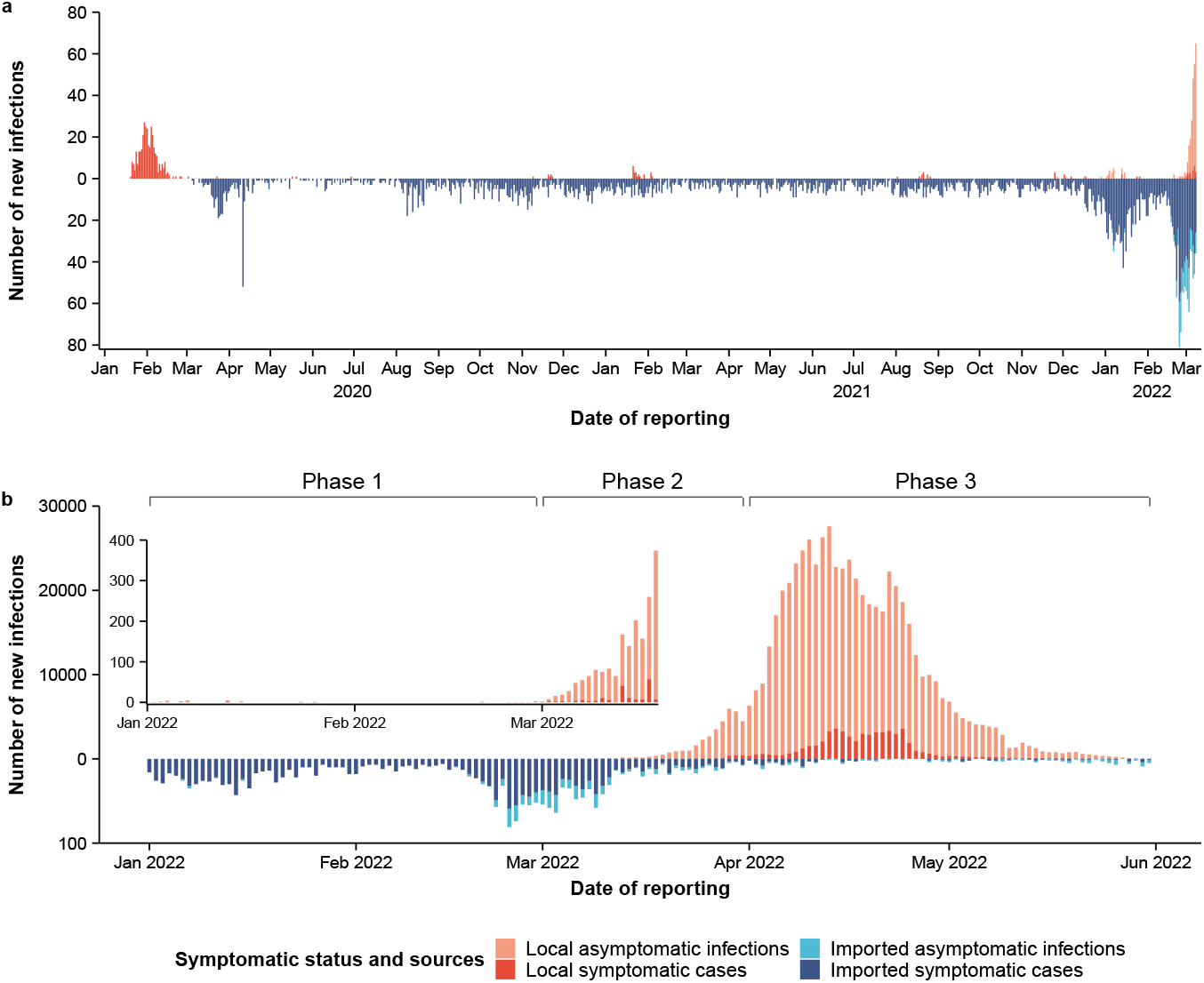
Temporal dynamics of local and imported SARS-CoV-2 infections in Shanghai since early 2020. **(a)** Number of reported SARS-CoV-2 infections in Shanghai between 2020-2022, stratified by local and imported infections. **(b)** The same as in (a), but for the period from January 1 to May 31, 2022.

Starting in mid-February 2022, Shanghai faced a significant surge of imported infections, the majority of which were linked to the Omicron wave in Hong Kong (**Supplementary Figure 3**). The number of imported infections reached a peak of 81 per day on February 24—more than 19 times the prior daily average in 2020-2021 (4.1 imported infections per day). Overall, from February 15 to March 15, a total of 1,112 imported infections were reported (**Figure 2b**). Compared to any other time point during the rest of the pandemic, the test positive rate among inbound travelers was at its highest throughout February 2022, reaching a peak of 96.3 infections per 100,000 travelers (**Supplementary Figure 4**).

The sudden upsurge of infected travelers ultimately led to a breach of the entry control program, which might have resulted in the cryptic transmission of the Omicron variant since early March. A cluster of 14 SARS-CoV-2 positive individuals was detected starting from March 1 among attendees of a square dance event for seniors. Because of the Omicron variant’s high transmissibility and immune evasive properties, as well as the delayed discovery of the outbreak, the contact tracing program was quickly overwhelmed, leading to a fast-growing epidemic that resulted in a total of 626,000 SARS-CoV-2 infections as of May 31, 2022 (**Supplementary Figure 5**).

Most infections (96.0%) were identified in individuals under quarantine (**Supplementary Figure 6a**). The extent to which the infections were identified by community screening (e.g., essential workers) gradually declined from 13.6% in Phase 2 to 2.7% in Phase 3. Meanwhile, the proportion of infections presenting with symptoms varied from 15.5% in Phase 1 to 9.8% in Phase 3 (**Supplementary Figure 6b**).

The Omicron outbreak was widespread throughout the entire area of Shanghai, with 99.5% (215/216) of the subdistricts reporting positive individuals. However, the spatial distribution of infections was highly heterogeneous, with 40% of the city area accounting for 80% of all infections; 35.5% of infections were identified in the Pudong New Area alone (**Supplementary Figure 5**). We divided the Omicron wave into three phases: Phase 1 (from January 1 to February 28), characterized by a high risk of importation; Phase 2 (from March 1 to 31), characterized by local community transmission; Phase 3 (after April 1), characterized by high levels of local transmission and the implementation of city-scale lockdowns to contain the outbreak. The total incidence of reported infections increased from 0.02 infections per 1,000 individuals in Phase 1 to 2.9 infections per 1,000 individuals in Phase 2 and reaching 21.9 infections per 1,000 individuals in Phase 3 (**Figure 3a-c**). Overall, an incidence of 24.8 infections per 1,000 individuals was observed (**Figure 3d**).

**Figure 3.**
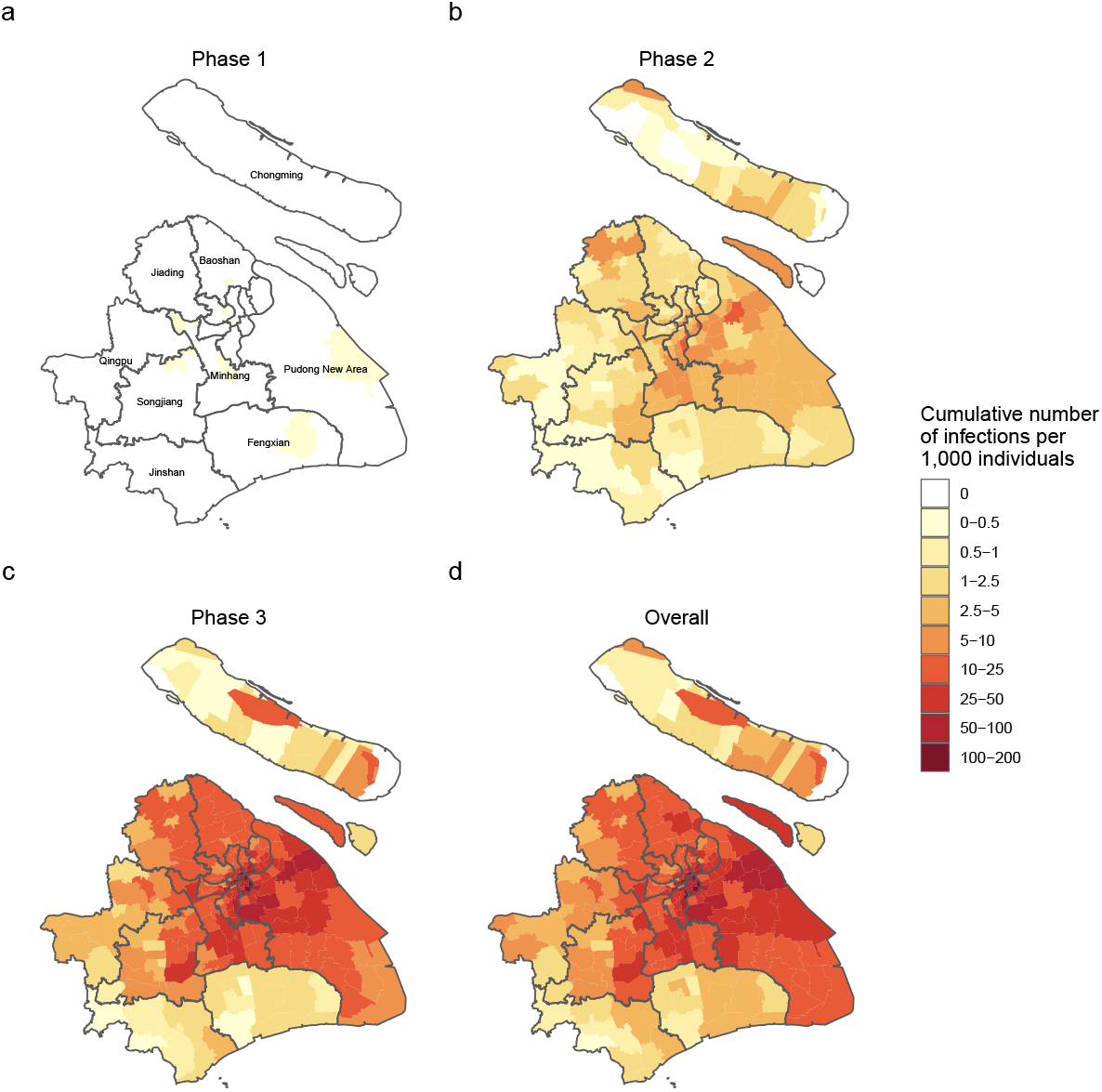
Geographical distribution of SARS-CoV-2 infections. **(a-d)** Cumulative number of new SARS-CoV-2 infections per 1,000 individuals in each phase and overall.

The spatial spread of the reported infections shows a clear trend from the city center towards adjacent suburban and rural areas (**Figure 4a-c**). Despite the implementation of a citywide lockdown in Phase 3, the outbreak continued to expand towards suburban and rural areas (**Figure 4c**). We estimated that spread speed of the infection throughout the city progressively slowed from an average of 544 meters/day in the first week of the outbreak (February 27-March 5) to approximately 325 meters/day at the end of March (March 25-31), before the city-scale lockdown (**Figure 4d**). Although this decreasing trend was common to all districts, the central areas (including the districts of Jing’an, Yangpu, Hongkou, Putuo, Changning, Xuhui, and Huangpu) showed the fastest rate of spread, especially in the early stage of the outbreak (**Supplementary Figure 7**).

**Figure 4.**
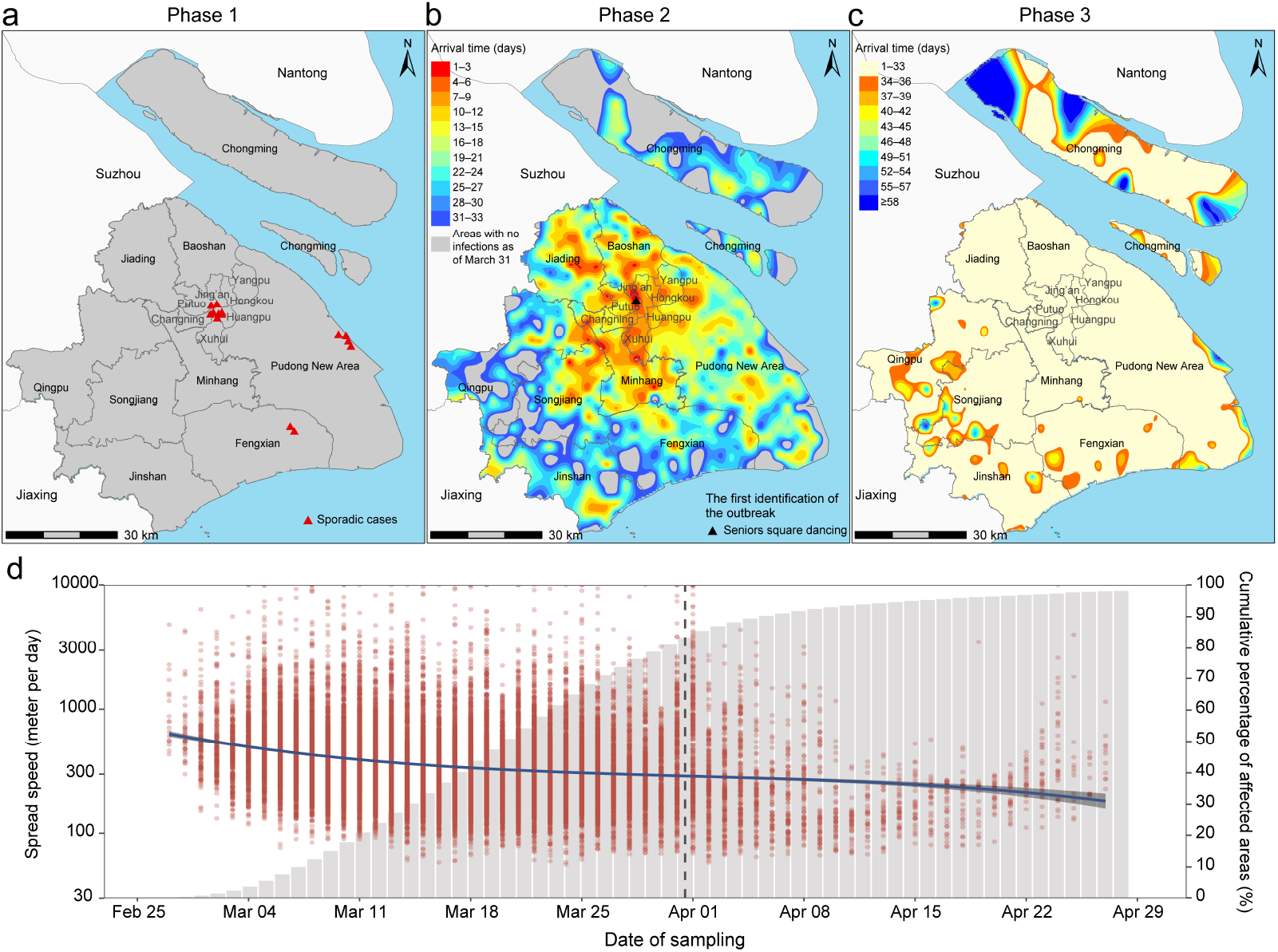
Spatial trends and speed of spread of the epidemic in the three phases. **(a)** Spatial location of the reported infections during the first phase of the epidemic. **(b-c)** Estimated arrival time of the epidemic in the different areas of Shanghai. Estimates are based on the thin spline regression of the interval between the time of the detection of the first infection in each 3 km × 3 km grid and February 27, 2022. Triangles indicate the potential source of the outbreak. **(d)** Estimated speed of spread of SARS-CoV-2 (left axis) and cumulative fraction of affected areas of Shanghai (right axis). Red dots indicate the speed of spread over time in each cell. The blue line indicates the average speed per day as obtained using a polynomial regression. Central areas contain the districts of Jing’an, Yangpu, Hongkou, Putuo, Changning, Xuhui, and Huangpu.

From March 16 to March 27, prior to the blanket lockdown of the entire city, Shanghai launched a set of targeted interventions and management strategies on a smaller geographical scale. The policy was adopted at the subdistrict level, and subdistricts were divided into high-risk areas (see definition in the **Methods** section), where the interventions were implemented, and non-high-risk areas. We further divided the non-high-risk areas into two categories based on their spatial proximity to the high-risk areas: moderate-risk areas (i.e., areas that were not classified as high-risk but were adjacent to high-risk areas) and low-risk areas (i.e., areas that were neither classified as high-risk nor adjacent to high-risk areas) (**Figure 5a**). From March 16 to March 29, the cumulative number of infections per 1,000 individuals was highest in high-risk areas (2.20, 95% CI, 2.18-2.22), followed by moderate-risk areas (1.51, 95% CI, 1.49-1.54) and low-risk areas (0.53, 95% CI, 0.49-0.57) (**Figure 5b-d**). The implemented targeted intervention strategies prior to the citywide lockdown were not sufficient to prevent infections to rise (**Figure 5e**), and *R*_*t*_ remained well above 1 (the epidemic threshold) across all three area categories for the entire period (**Figure 5f**). Moreover, *R*_*t*_ in moderate-risk and low-risk areas increased slightly from March 20 to March 24. The sensitivity analysis that considered alternative values for the delay between sampling and reporting showed similar patterns (**Supplementary Figure 8**). Overall, the growth rate of the epidemic in Phase 2 was estimated to be 0.216 per day (95% CI, 0.210 to 0.222), with infections doubling every 3.21 days (95% CI, 3.12 to 3.29) (**Figure 5g**).

**Figure 5.**
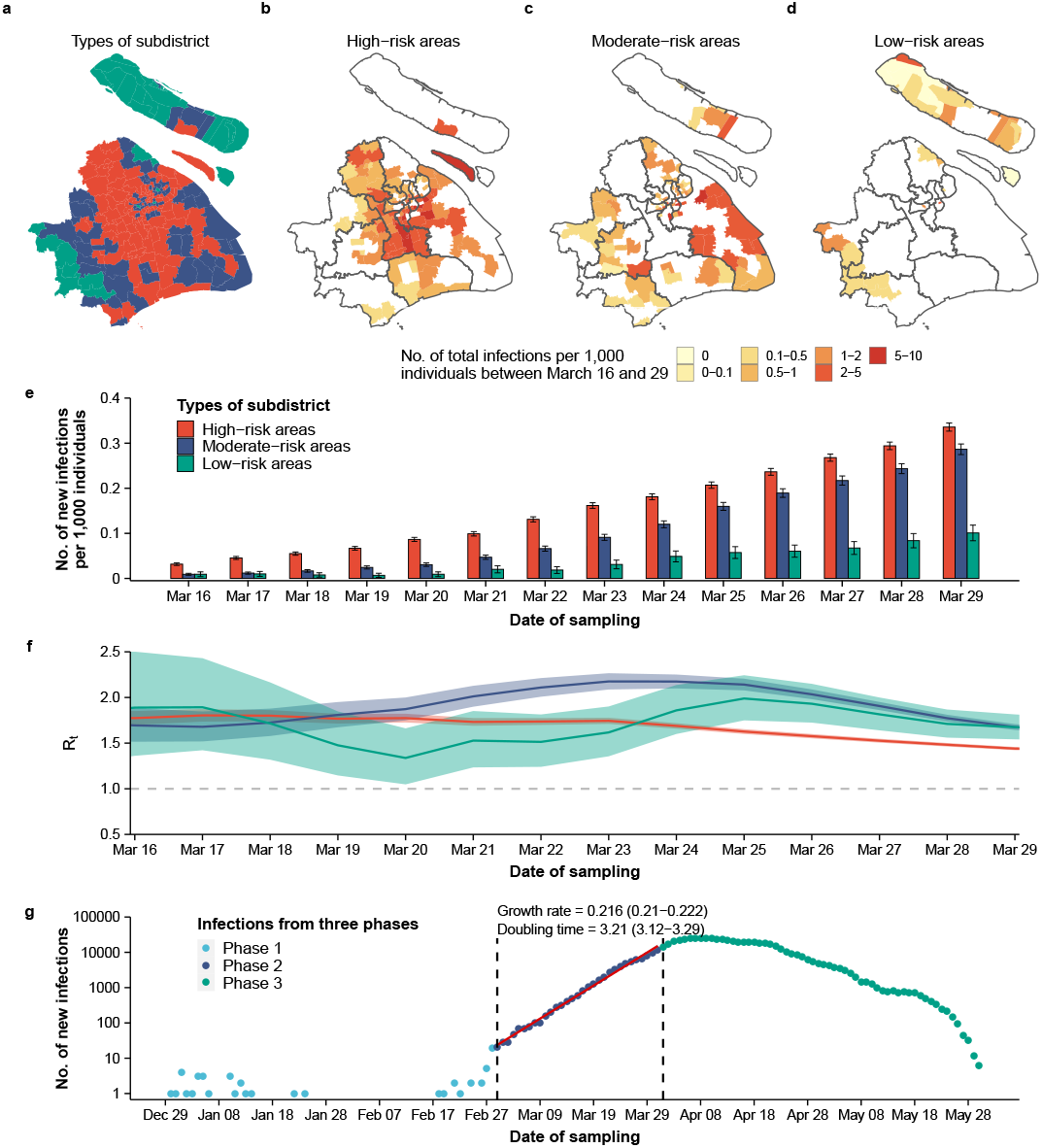
Characterization of the epidemic dynamics between March 16 and March 29, 2022. (a) Location of high-risk, moderate-risk, and low-risk areas. For each area, its highest risk classification was used. (b-d) Number of reported infections between March 16 and March 29 by area type. (e) Number of new reported infection per 1,000 individuals by area type. (f) Estimated *R*_*t*_ between March 16 and March 29 by area type. (g) Estimated epidemic growth rate and doubling time (days).

Three days after the implementation of a citywide lockdown on April 1 (Phase 3), *R*_*t*_ started to decrease and fell below the epidemic threshold on April 14 (**Figure 6a-b**). Despite the marked differences in *R*_*t*_ observed between eastern and western Shanghai when the targeted interventions were adopted (Phase 2), immediately after the implementation of the citywide lockdown, the same level of SARS-CoV-2 transmission (*R*_*t*_) was observed in all of Shanghai (**Figure 6b** and **Supplementary Figure 9**, where a sensitivity analysis confirming this finding is presented).

**Figure 6.**
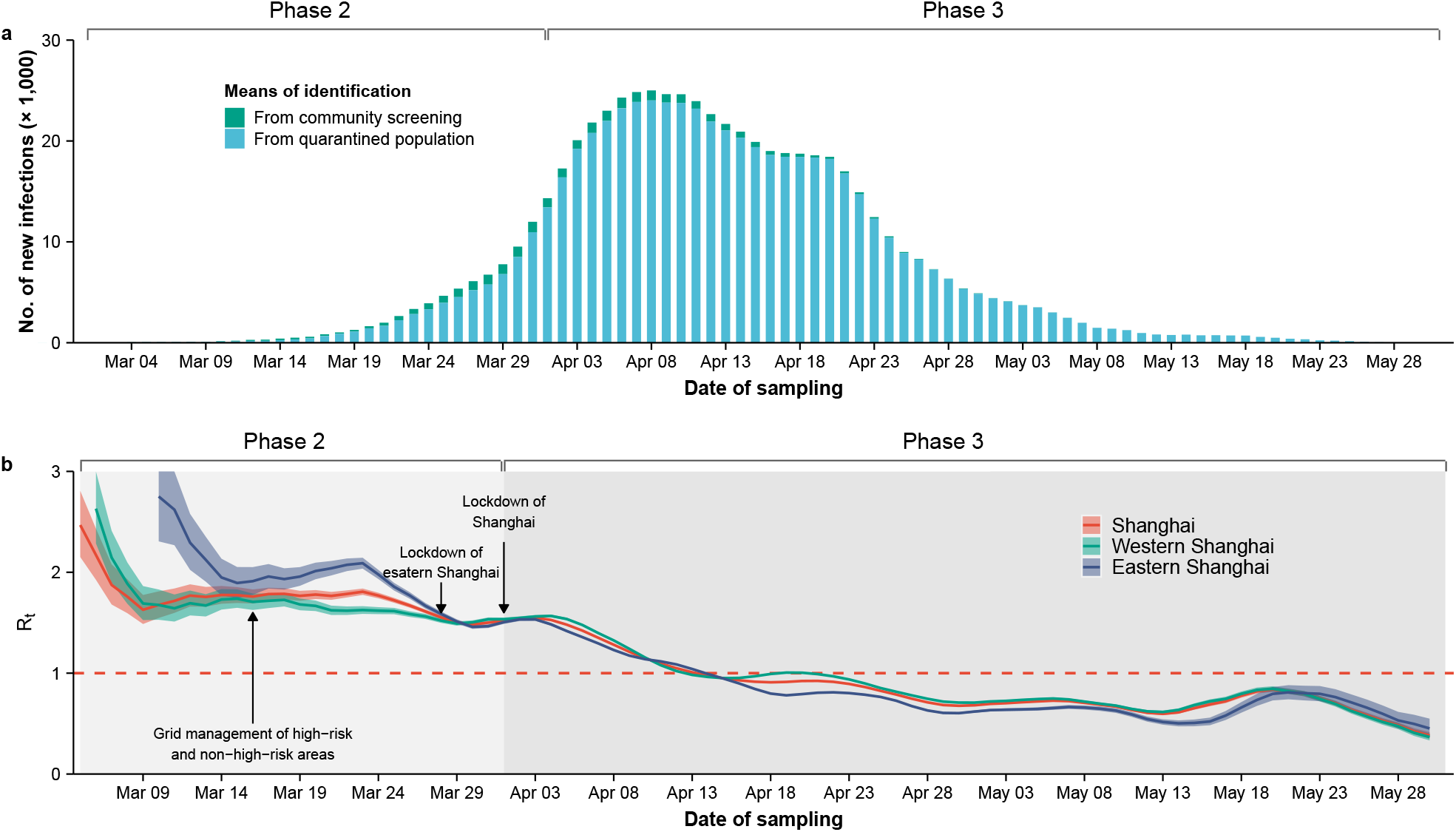
Epidemic dynamics under the effect of interventions. **(a)** Number of new SARS-CoV-2 infections by date of sample collection for means of identification. **(b)** Estimated *R*_*t*_ (mean and 50% confidence interval) in eastern, western, and all Shanghai areas.

## Discussion

This study provides a comprehensive picture of the spatiotemporal dynamics of the Omicron outbreak in Shanghai in 2022. Our findings highlight that, given the Shanghai population’s susceptibility to the SARS-CoV-2 Omicron BA.2 variant and its high transmissibility, the heightened importation risk of infected individuals from areas with widespread viral circulation was capable of triggering a major epidemic wave. The targeted interventions implemented in March were not enough to halt transmission. The citywide lockdown imposed on April 1 led to a decrease in transmission, bringing the reproduction number below the epidemic threshold in the entire city. Overall, the Omicron outbreak in Shanghai was successfully contained through a combination of stringent measures.

Our findings suggest that when facing a high influx volume of travelers from high-risk areas, breaches in the screening and quarantine of travelers may have the potential to trigger major outbreaks. It is also possible that the wave was simultaneously triggered by domestic travelers, as suggested by previous field investigations by local authorities.^23^ We cannot exclude that the outbreak might have originated from multiple sources that led to simultaneous transmission chains prior to the identification of the first local transmission event.

Compared to previous local SARS-CoV-2 outbreaks that emerged in China after the original COVID-19 wave, the rapid spread of the Omicron outbreak in Shanghai was likely driven by the high transmissibility and/or immune-escape properties of Omicron BA.2.^24^ The immunological landscape of Shanghai at the onset of this outbreak was fragile, with essentially no prior natural immunity (less than 0.01% of the population was infected prior to the outbreak^25^). In addition, although the coverage of the primary vaccination series exceeded 90% of the total population, the vaccination coverage for individuals aged ≥ 60 years was relatively low (62% for the primary series and 38% for the booster dose^26^). Moreover, the inactivated vaccines used in Shanghai provided very low protection against Omicron variant infection (approximately 17.0% after receiving a booster dose^11^, based on the level of neutralizing titer), which quickly wanes over time.^27^ As a result, the Shanghai population was particularly vulnerable to the Omicron variant. Nonetheless, inactivated vaccines could provide high protection against severe outcomes.^9,28^ These considerations further emphasize the importance of administering vaccines, especially to the most vulnerable segments of the population, and deploying immunization stratgies with improved effectiveness against the current circulating variants through procuring currently available mRNA vaccines, approve other domestically developed vaccine candidates with borader protection against Omicron and future variants, and heterologous boosting strategies that have demonstrated improved antibody response.

Another factor that led to the major outbreak might have been the large number of asymptomatic carriers that contributed to widespread silent transmission.^29^ Here, we found a high proportion (more than 90%) of asymptomatic infections, larger than that estimated for the ancestral lineage (69%^30^), but in line with other estimates for other SARS-CoV-2 variants in the presence of vaccination (85%^31^). Lower proportions of asymptomatic infections were reported in previous studies,^32^ including for the Omicron BA.2 outbreak in Hong Kong^19^; however, those estimates were obtained in the absence of repeated screenings of the population. Moreover, the criteria adopted for the definition of asymptomatic infection may vary across locations and the study period.

Between March 1 and March 31, a series of public health measures were adopted to reduce transmission between individuals living in areas at different risk levels, and the estimated doubling time in this phase of the epidemic was comparable to that of the Omicron wave in Hong Kong (3.2 days as compared to 3.4 days estimated for Hong Kong^19^). However, the implemented interventions were insufficient to curtail the epidemic. In fact, the repeated two-day lockdowns and population screenings implemented in high-risk areas were not sufficient to halt SARS-CoV-2 transmission, and these failed to identify all infected individuals (e.g., those in the early course of the infection, when the viral load was below the detection limit^33^). Once the two-day local lockdowns were lifted, unidentified infected individuals were allowed to freely move and transmit the infection to other areas, as shown by the rise in new daily infections identified in areas adjacent to high-risk areas.

The implementation of the citywide lockdown prevented further growth of the epidemic and successfully contained the outbreak. However, given the number of infectious individuals at the time of implementation (over 10,000 new infections reported per day), it took 13 days for *R*_*t*_ to fall below the threshold. It is also possible that it was difficult to transport and isolate all positive individuals in a timely manner into dedicated facilities during the lockdown. Moreover, residents may still have been exposed to the virus inside their buildings.^34^ It is thus important to carry out quantitative evaluations of the unavoidable risks posed by the adopted strategies. Empirical evidence from Shanghai shows that it is possible to maintain containment policies against the highly transmissible Omicron BA.2 variant, although this required unprecedented efforts to achieve. However, it is important to stress that the interruption of onward transmission in Shanghai might have prevented the infection from spreading into other cities, potentially preventing a major public health crisis in mainland China.^11^ Nonetheless, it is important to stress that the measures adopted during the Omicron outbreak in Shanghai, including a strict and prolonged city-scale lockdown, would be socially costly and impractical in the long term, emphasizing the importance refining NPI stratgeties to be less disruptive of daily lives and boosting population immunity level to significantly reduce the morbidity and motality burden of SARS-CoV-2.^11^

A strength of our study is the unique features of the analyzed dataset. In fact, Shanghai’s population-based screening policy, which allowed infections to be identified, likely resulted in a high infection ascertainment rate and the identification of the majority of infections throughout the outbreak. This allowed us to investigate the spatiotemporal spread of the Omicron BA.2 variant as well as the impact of the implemented interventions, all without significant amounts of underreporting from surveillance systems, as experienced in other areas. However, our study suffers from the limitations rooted in the uncertainty and fragmentary nature of publicly available sources. First, key variables, such as the date of symptom onset and the addresses of infected individuals, suffered from a high level of missing data. Second, the sparse information on the demographic characteristics of positive individuals limits the scope of our study. Third, the final clinical outcome of infected individuals was only partially available (e.g., we had no information about patients requiring intensive care treatment). Moreover, given the real-time nature of this study and the censoring of the final outcome data, an analysis of the COVID-19 burden is not possible at this point. Finally, this study does not provide quantitative estimates of the impact of specific interventions on transmission dynamics, but providing an overall assessment of the synergetic effect of the adopted interventions.

In conclusion, this study provides a quantitative description of the spatiotemporal spread of the Omicron BA.2 variant in Shanghai and explores the impact of the multifaceted implemented response measures. Our findings highlight the risk of widespread outbreaks in mainland China, particularly under the large pressure of imported infections. The successful containment of the Shanghai outbreak through the implementation of a strict and prolonged citywide lockdown shows that it is possible to successfully contain an Omicron outbreak in China, although it requires the implementation of a set of very stringent interventions. Disentangling the effects of each of the performed interventions might provide further insights into key public health priorities when faced with the emergence of future novel variants.

## Data Availability

The data and code that support the findings of this study will be made available in GitHub upon manuscript acceptance

## Contributors

H.Y. conceived and designed the study. H.Y. and JJ.Z. supervised the study. X.D., JJ.Z., Z.C., JY.Z., J.C., Y.W., Y.T., N.Z., X.Y., R.S., X.X., X.Z., S.G., Y.L., and L.Y. collected and checked data. Z.C., JJ.Z., L.F., T.C., YP.W., JY.Z., J.C., H.L., K.S., Y.W., T.W., and Y.T. analyzed the data. Z.C., JJ.Z.. and H.Y. wrote the first draft of the manuscript. Z.C. JJ.Z, K.S., M.A., L.F., J.Y., and H.Y. interpreted the results and revised the content critically. All authors approved the final version for submission and agreed to be accountable for all aspects of the work.

## Acknowledgments

The findings and conclusions in this report are those of the authors and do not necessarily represent the official position of the NIH. This study was supported by grants from the Key Program of the National Natural Science Foundation of China (grant 82130093 to H.Y.). The funders had no role in study design, data collection, data analysis, data interpretation or writing of the report.

