## Appendix for "Epidemiological characteristics and transmission dynamics of the outbreak caused by the SARS-CoV-2 Omicron variant in Shanghai, China: a descriptive study"

### Supplementary methods

#### Case definition

Laboratory-confirmed cases were categorized into four types based on clinical severity: mild, moderate, severe, and critical. According to the updated guidelines on the diagnosis and treatment of patients with COVID-19 (ninth version),^1^ mild cases are defined as those with mild symptoms such as fever, fatigue, and loss of taste/smell but without radiographic evidence of pneumonia. Moderate cases refer to those with typical symptoms of respiratory infection (e.g. fever, dry cough, fatigue) and radiographic evidence of pneumonia. Severe cases refer to those with any one of the following: breathing problems, low oxygen saturation, low PaO_2_/FiO_2_, or progressive symptoms combined with pulmonary imaging showing obvious progression of lesions (> 50%) within 24–48 hours. Critical cases refer to patients who meet any one of the following three criteria: respiratory failure, shock, or organ failure requiring intensive care unit admission.

#### Case surveillance and detection

In addition to the routine case surveillance network (detailed description available in reference^2^), several rounds of risk-based or population-based molecular test screenings were carried out in Shanghai, allowing for the early detection of SARS-CoV-2-infected individuals and the timely implementation of interventions. Moreover, self-performed rapid antigen test screenings were performed as a supplement to nucleic acid tests in regions with insufficient testing capacity. Any positive antigen test result required confirmation by a nucleic acid test.

#### Management of infections and cases

The protocols adopted in Shanghai prescribe that all asymptomatic infections and mild cases need to be isolated in makeshift isolation hospitals to prevent further community transmission, except for vulnerable groups (e.g. children, pregnant women, haemodialysis, and oncological patients), who are required to be admitted to designated hospitals with SARS-CoV-2 treatment capacity (hereafter referred to as COVID-19 treatment hospitals). Moderate, severe, and critical COVID-19 cases were also admitted to COVID-19 treatment hospitals.

Patients in makeshift isolation hospitals were discharged if two consecutive nucleic acid test results (sampled at least 24 h apart) were negative. For patients in COVID-19 treatment hospitals, in addition to the criterion of two consecutive negative results, the following criteria were adopted: normal body temperature for more than three days and significant improvement of respiratory symptoms and lesions shown on pulmonary imaging.^1^

#### Data collection

The city of Shanghai is divided into 16 districts and 216 subdistricts. Subdistrict-level population data after 2017 were derived from the Seventh National Census of China and the latest reports by local authorities^3^. For the 67 subdistricts with unavailable population data after 2017, the subdistrict-level population data for 2020 were inferred from the population size of each district in 2020 and the population proportion of each subdistrict in the Sixth National Census in 2010.

The monthly count of inbound travelers to Shanghai for the period from March 2020 to February 2022 was downloaded from the Official Aviation Guide (OAG) database. Administrative boundaries in Shanghai were downloaded from Amap (a Chinese web-based service providing geographical information, <https://ditu.amap.com/>); boundaries were updated to reflect recent administrative changes. All datasets were cross-checked by two co-authors, and in cases of unmatching data, the priority was given to those from the original data sources.

#### Inference of delay between sampling and reporting

The daily number of specimens between March 16 and April 27 was collected from public sources, and missing data were estimated by linear extrapolation based on the time series of cumulative specimens. We regarded the maximum lag to be statistically significant as the mean delay. Cross-correlation analysis was performed for two periods (March 16 to April 3; April 4 to April 27), the division of which was based on the start of the citywide screening protocol on April 4.

We assumed that the time interval between sampling and reporting was 2 days before March 15, given that no massive screening was performed over that period.^4^ A lag of 3 days (ρ = 0.46, *p* < 0.05; ρ = 0.51, *p* < 0.05) was estimated for both periods (**Supplementary** **Figure 2c–d**), and the delay for the period between April 28 and May 14 was also assumed to be 3 days. The sampling date was inferred by generating a random value from a gamma distribution with a mean of three days and subtracting it from the reporting date.^2^ In the sensitivity analysis, we considered a four-day delay for the periods from March 16 to May 14. The daily nucleic acid detection capacity has grown to 8.5 million and the daily reported number of SARS-CoV-2 infections was less than 1,000 in May 15; thus, we assumed that the time interval between sampling and reporting was 2 days after May 15, similar to the delay before March 15.

#### Sampling of individual addresses

Detailed address information for each infected individual was released before March 18. Afterwards, the list of addresses for all daily new infections was reported by district at the street block level, but we could not access the exact location of each individual. The specific location for each individual was sampled from the list of addresses that reported infections that day by district. The subdistrict where the individuals were located were then obtained based on the sampled addresses.

#### Progression of pre-symptomatic infections

If a symptomatic infection was detected before symptom onset (i.e. during the incubation period), it would be initially reported again as an asymptomatic infection. However, once they developed symptoms, they were re-characterized as symptomatic. Therefore, for each symptomatic case, we performed the following de-duplication procedure. The infection time of each symptomatic case was inferred based on a gamma-distributed incubation period of 4.4 days on average.^19^ Then, based on the infection date, we matched the record with the previous asymptomatic infections at the same address. Finally, we revised the symptomatic status of the matched asymptomatic infections into symptomatic cases and subtracted them from the asymptomatic population accordingly.

### Supplementary results

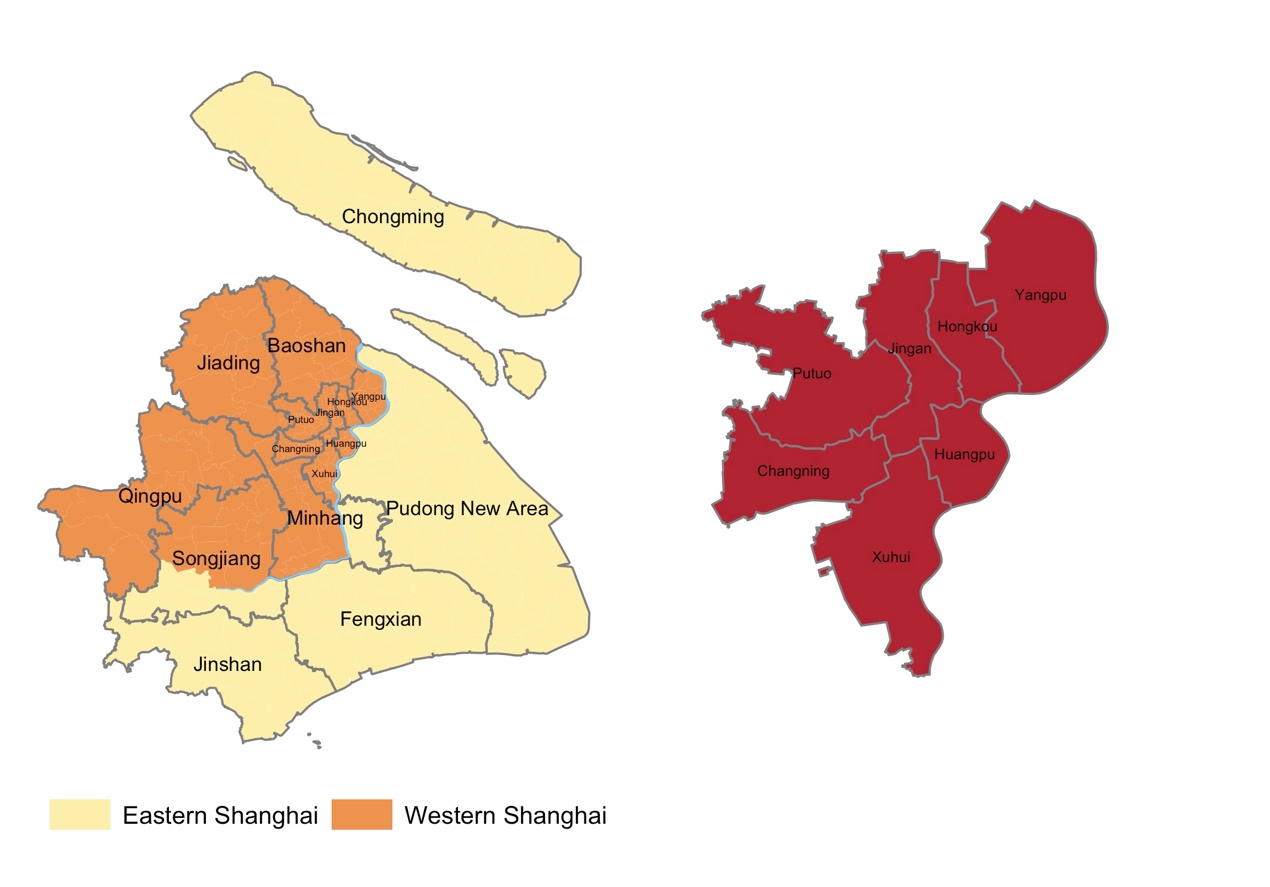

#### Supplementary Figure 1. Geographic division of eastern and western Shanghai.

Eastern and western Shanghai are naturally separated by the Huangpu River (blue layer). Specifically, eastern Shanghai contained the Pudong New District, Fengxian District, Jinshan District, Chongming District, as well as partial Minhang and Songjaing District; while the rest areas were grouped into western Shanghai. The right panel shows the central areas of Shanghai.

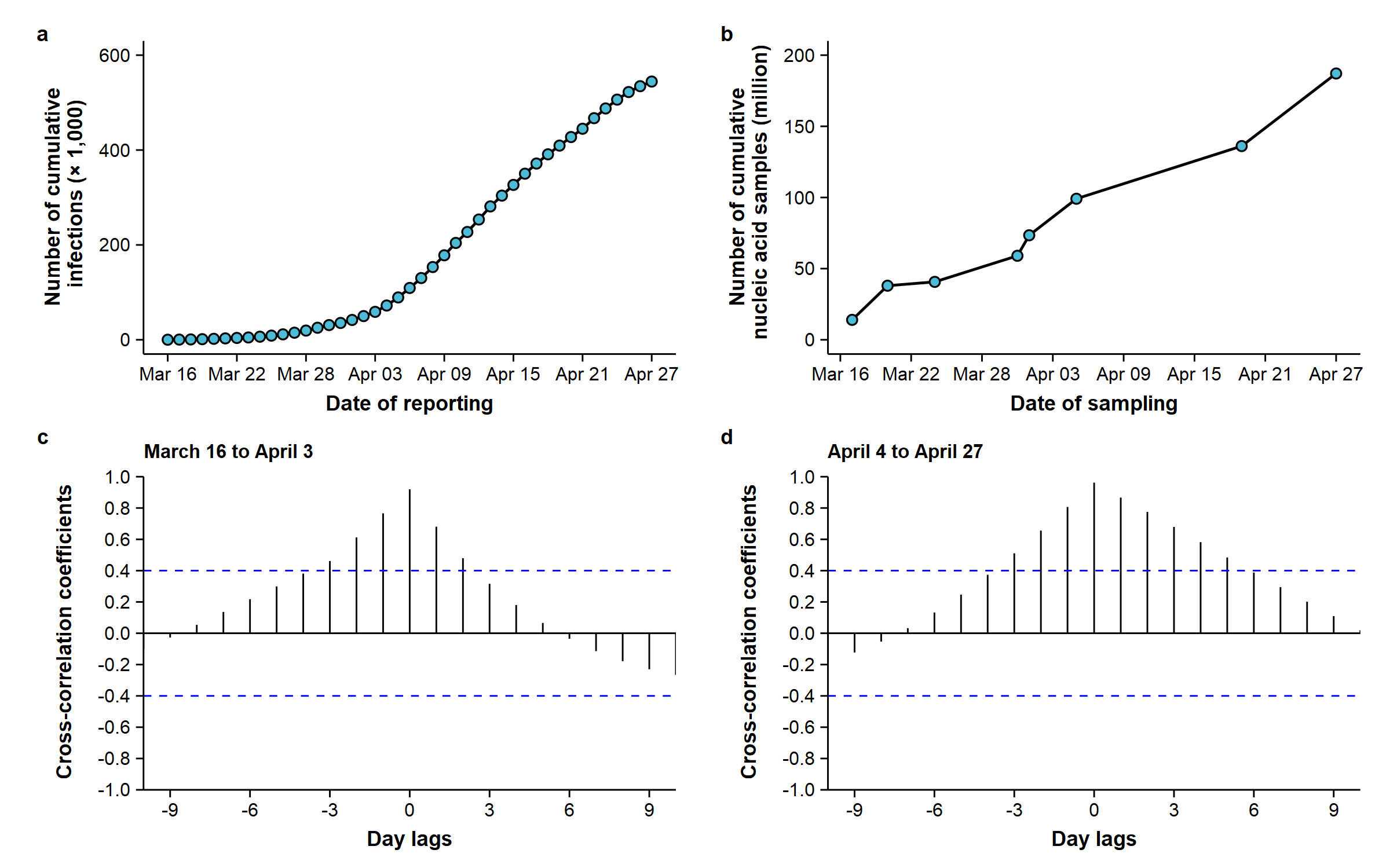

#### Supplementary Figure 2. Analysis of cross-correlation coefficients to infer the delay between sampling and reporting.

(a) Cumulative number of SARS-CoV-2 infections in Shanghai by date of reporting. (b) Number of SARS-CoV-2 nucleic acid samples accumulated by the eight major time points of nucleic acid sampling reported in Shanghai by date of sampling. (c-d) Cross-correlation coefficients between the cumulative number of reported SARS-CoV-2 infections and the cumulative number of sampled specimens under different lengths of lag in two periods (March 16 to April 3; April 4 to April 27, data on number of sampled specimens after April 28 was unavailable until now).

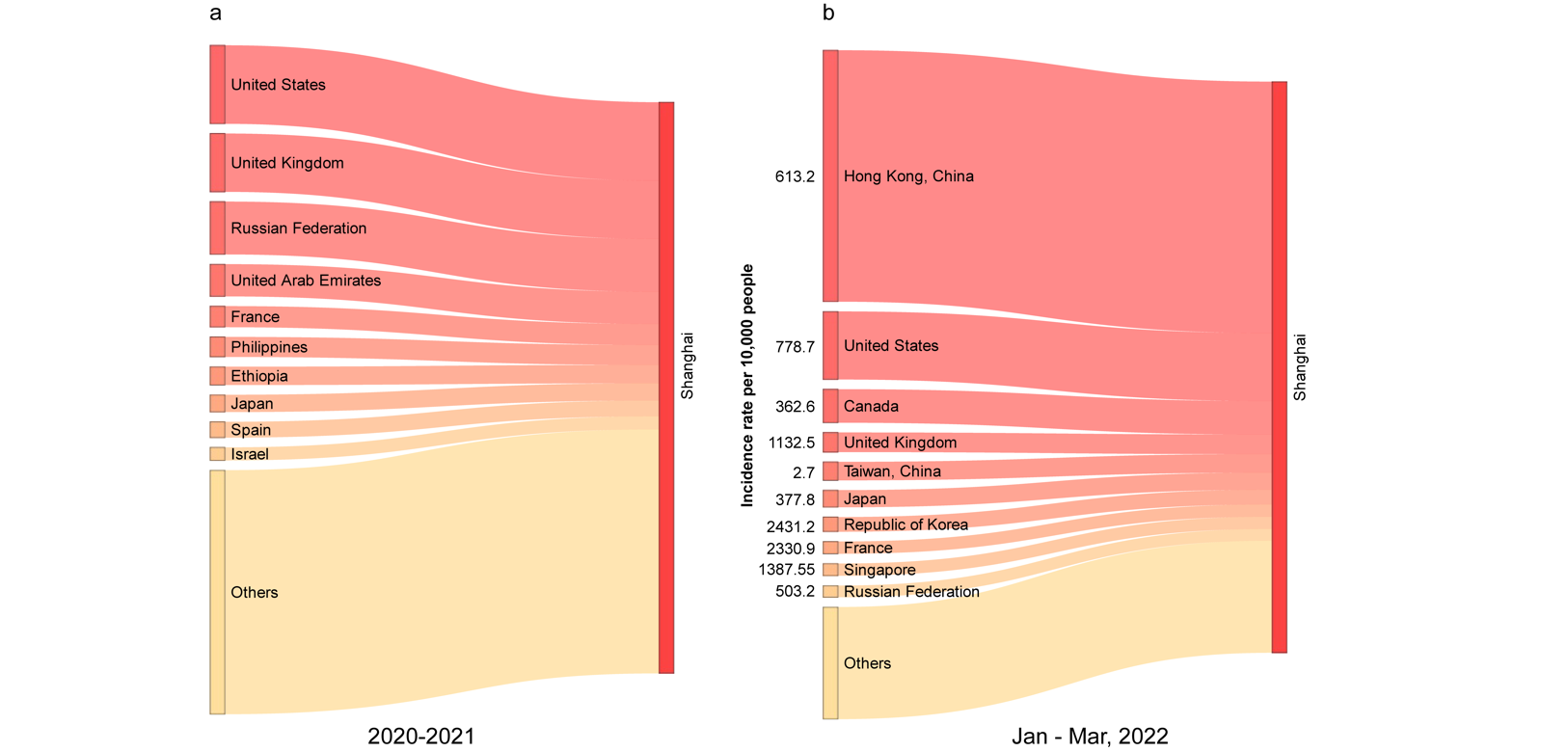

#### Supplementary Figure 3. The locations origin of imported infections into Shanghai.

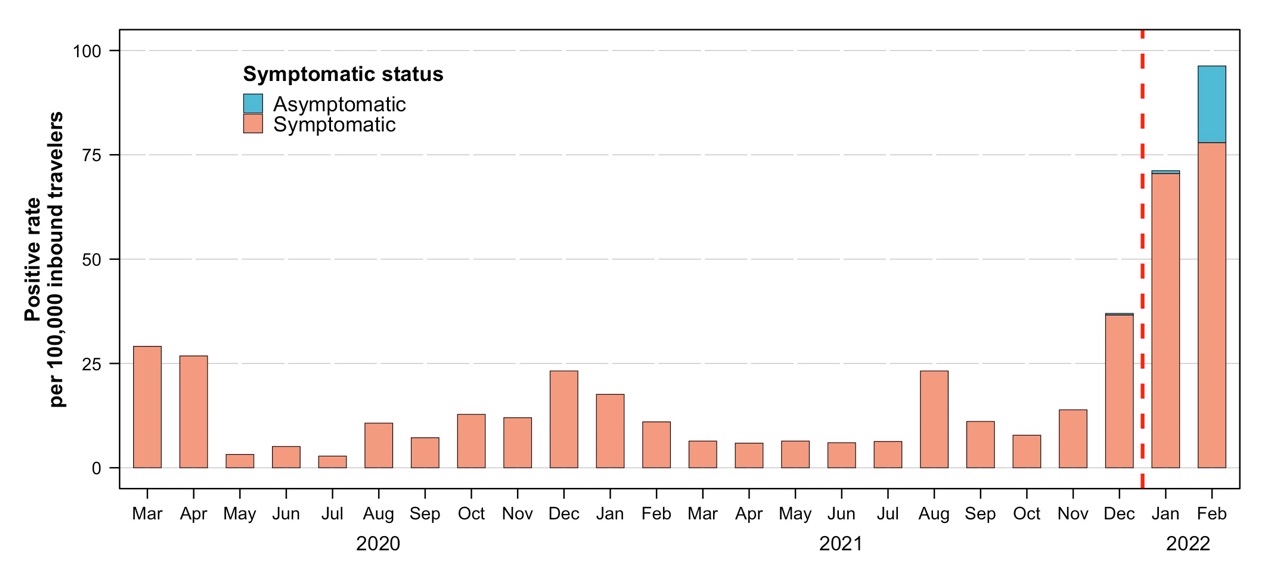

#### Supplementary Figure 4. The positive rate per 100,000 inbound travelers.

We restricted this analysis to those inbound travellers entering via international flights.

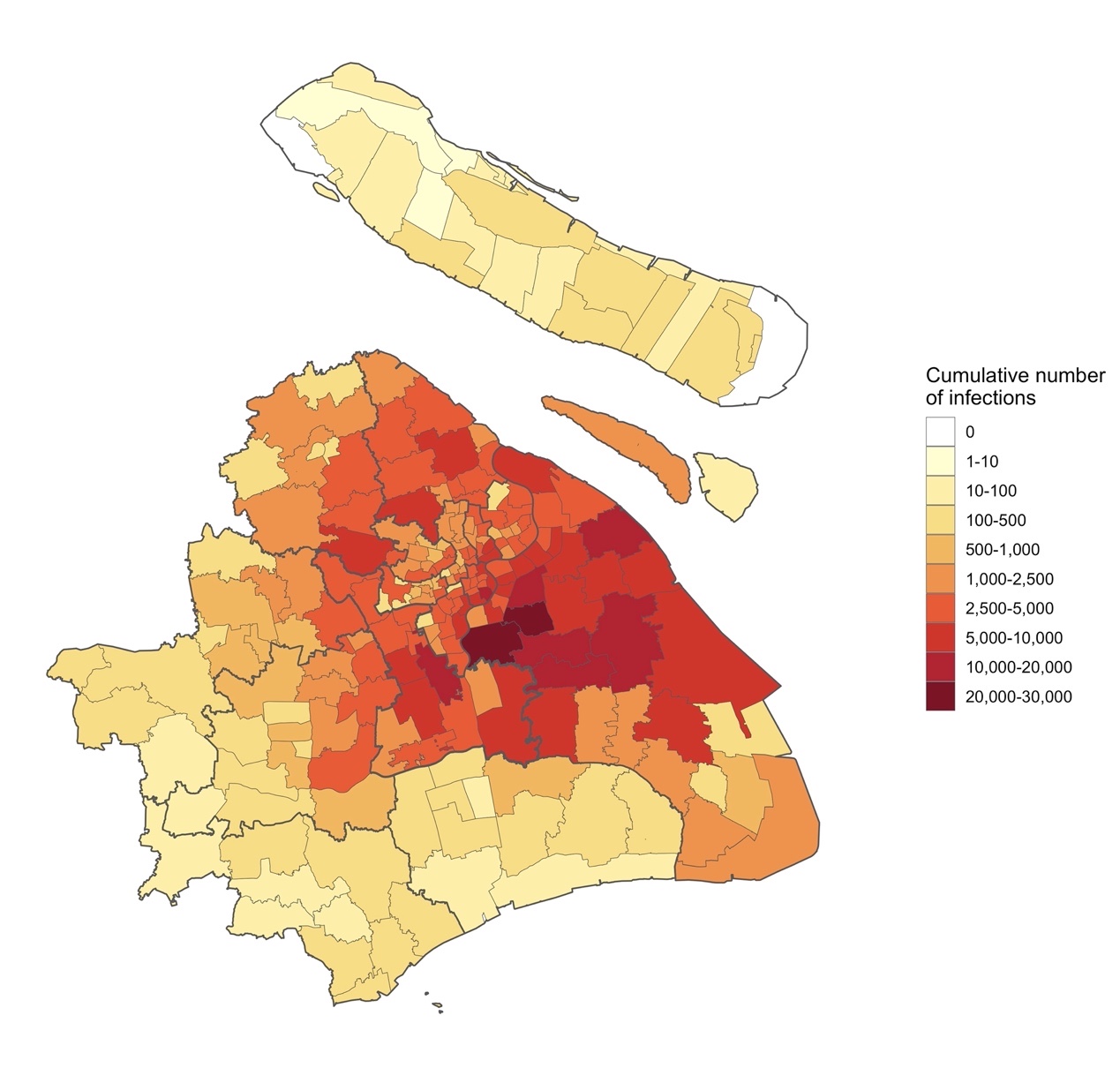

#### Supplementary Figure 5. Geographical distribution of cumulative SARS-CoV-2 infections since early 2022 in Shanghai.

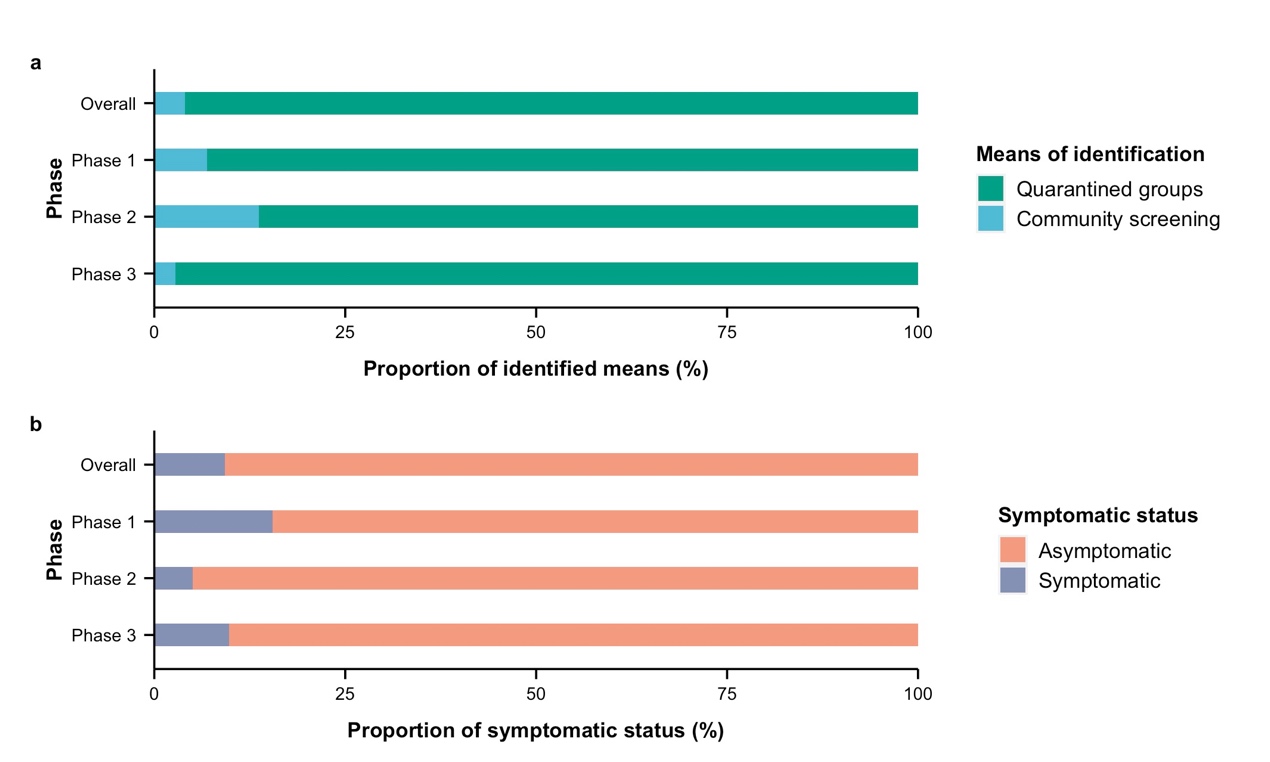

#### Supplementary Figure 6. Source of identification and symptomatic status of infections across the three phases.

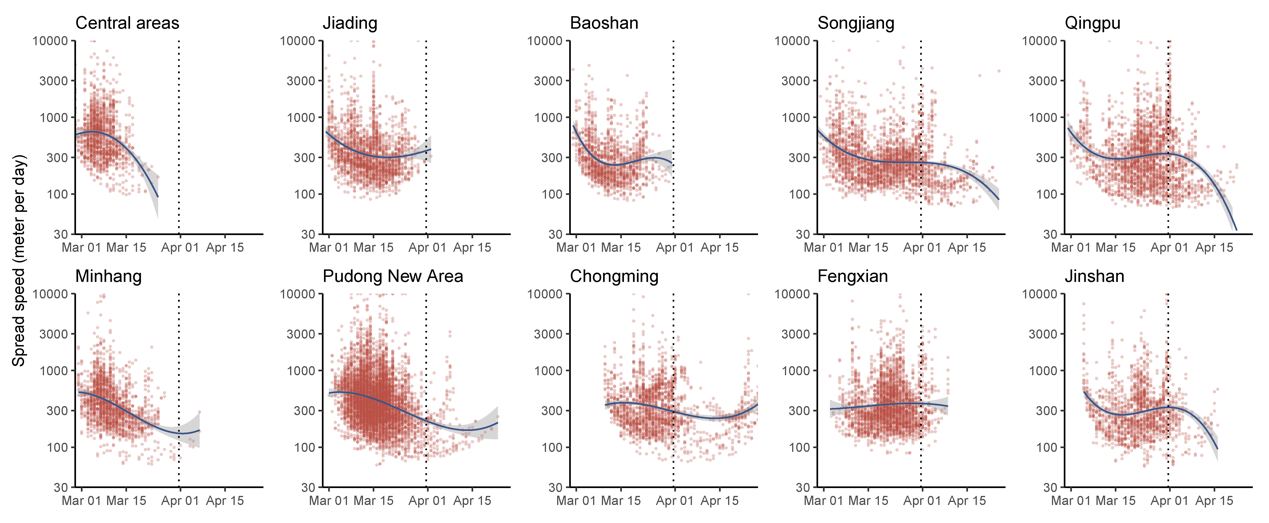

#### Supplementary Figure 7. The speed of spread of the epidemic by district.

Central areas contain the districts of Jing’an, Yangpu, Hongkou, Putuo, Changning, Xuhui, and Huangpu. Red dots indicate the speed of spread over time in each cell. The blue line indicates the average speed per day as obtained by using a polynomial regression.

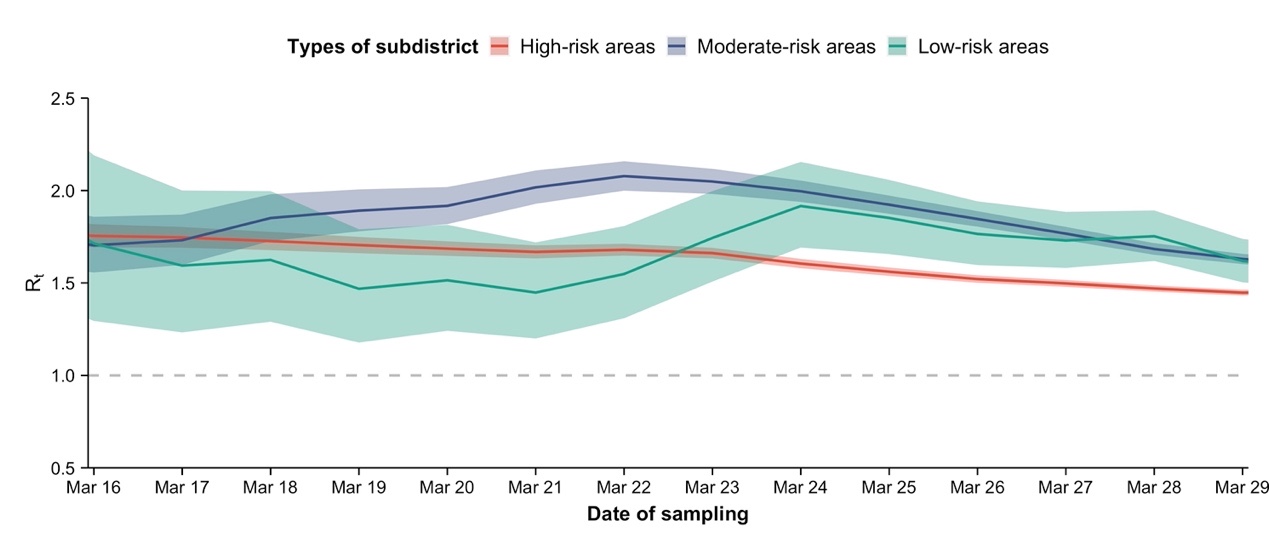

#### Supplementary Figure 8. Estimated *R_t_* between March 16 and March 29 by area type using a longer interval delay between sampling and reporting.

A gamma distribution of four-day delay was used for the periods from March 16 to May 14.

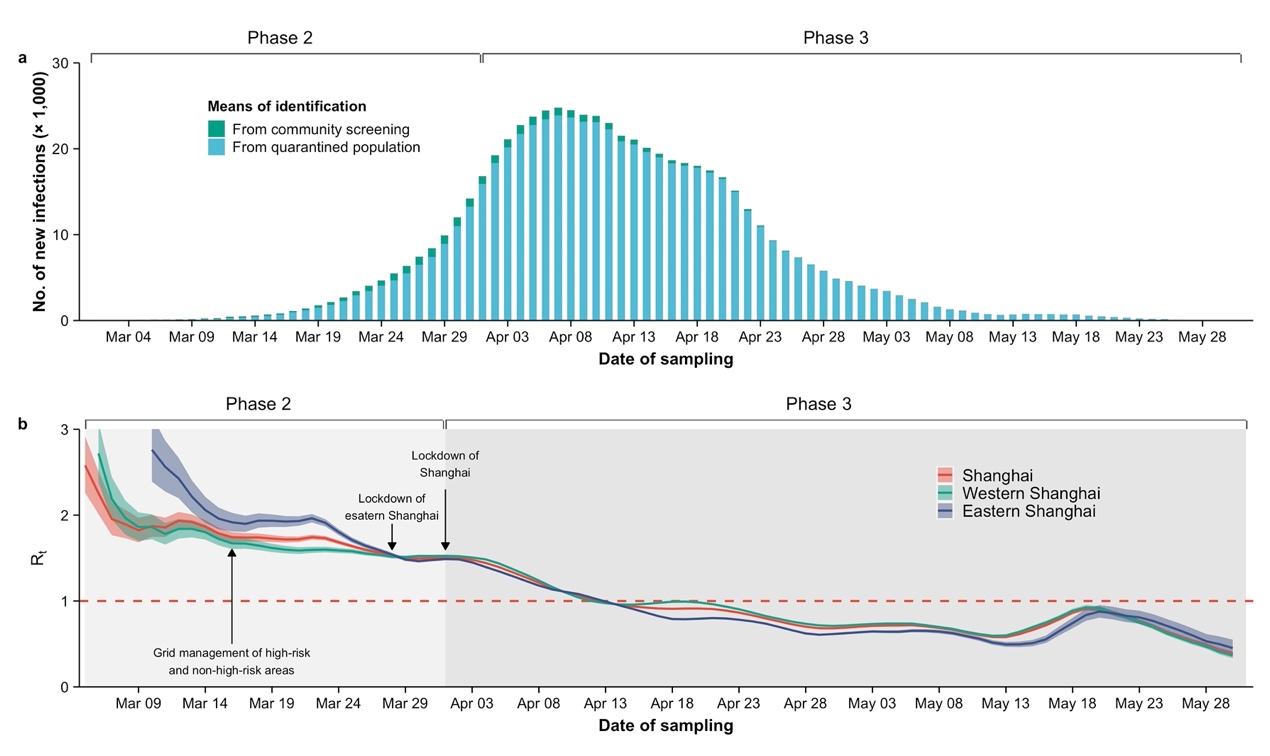

#### Supplementary Figure 9. Epidemic dynamics under the effect of interventions using a longer interval delay between sampling and reporting.

(a) Number of new SARS-CoV-2 infections by date of sample collection for means of identification. (b) Estimated *R_t_* (mean and 50% confidence interval) in eastern, western, and the entire Shanghai. A gamma distribution of four-day delay was used for the periods from March 16 to May 14.

#### Supplementary Table 1. Main publicly data sources.

| **Date types** | **Agency of sources** | **Links** |
| --- | --- | --- |
| Aggregated and individual infections | Shanghai Municipal Health Commission | https://wsjkw.sh.gov.cn/xwfb/index.html |
|  | Shanghai Municipal Government | https://www.shanghai.gov.cn/nw48546/index.html |
|  | Official Accounts of Shanghai | https://weibo.com/shanghaicity |
| Lists of high-risk area | Social media | <https://baijiahao.baidu.com/s?id=172747316>3527481719&wfr=spider&for=pc  <https://www.toutiao.com/w/1728024361311247/?app=news_article&timestamp>  =1647991687&use_new_style=1&wxshare_count=1&tt_from=weixin&utm_source  =weixin&utm_medium=toutiao_android&utm_campaign=client_share&share_token  =50ee15b1-a62e-49b6-a696-37cc50fed3a7  <https://mp.weixin.qq.com/s/isrnb_ljeQjz4rhyCgBg0A>  <https://mp.weixin.qq.com/s/2EochxcBH47gtWwVXRvk-w>  <https://mp.weixin.qq.com/s/k6HtOOCSR_Ahy2JjmqrVVA>  <https://mp.weixin.qq.com/s/g0mPvd1gsI0neCAWXXWSJg>  <https://mp.weixin.qq.com/s/1GJ6JaPv3Q51wEEdLOhSgw>  <https://mp.weixin.qq.com/s/yQVd1OqGdQcZ6VpKUeEVYw>  <https://mp.weixin.qq.com/s/gUD2qOvtfa5WyqyTOr_h2A>  <https://mp.weixin.qq.com/s/uc2K5S-NNvfjno4jueOvRw> |

#### Supplementary Table 2. Completeness assessment of variables in line lists.

| **Variable** | **Completeness of raw data (%)** | **Completeness after inference (%) ^a^** |
| --- | --- | --- |
| Address | 0.2 (1426/626840) | 99.7 (625228/626840) |
| District | 100.0 (626763/626840) | 100.0 (626763/626840) |
| Subdistrict | 0.2 (1426/626840) | 99.7 (625228/626840) |
| Symptomatic status | 100.0 (626840/626840) | 100.0 (626840/626840) |
| Source of identification | 100.0 (626840/626840) | 100.0 (626840/626840) |
| Reporting date | 100.0 (626840/626840) | 100.0 (626840/626840) |

^a^ Inference was only applied to variables of address and subdistrict.

#### Supplementary Table 3. The detailed NPI strategies performed in Shanghai since early 2022.

| **Intervention types** | **Administrative unit** | **Round** | **Implemented time point** | **Suspended time point** | **Details** | **Sources** |
| --- | --- | --- | --- | --- | --- | --- |
| **Part I. Baseline non-pharmaceutical interventions (NPIs)** | | | | | | |
| **Border control measures** | | | | | | |
| Negative certificate of nucleic acid | Shanghai | - | Before 2022 | Now | It depends on country of origin, and antibody tests or antigen tests might be also necessary. | <http://www.china-embassy.org/lsfw/sjc/202204/t20220415_10668659.htm>  https://www.mfa.gov.cn/ce/ceuk//chn/lsfw/lsxz/t1854637.htm |
| Quarantine for inbound travelers at designated facilities | Shanghai | - | Before 2022 | Now | 14 days of centralized isolation in designated hotels | http://sh.bendibao.com/news/20211212/246484.shtm |
| Home-quarantine | Shanghai as final destination | - | Before 2022 | Now | After 14 days of centralized isolation, those who have a fixed place of residence in Shanghai and meet the conditions for home health monitoring will undergo 7-day home health monitoring. | http://sh.bendibao.com/news/20211212/246484.shtm |
| Fuse measures for international scheduled passenger flights | Shanghai | 1 | 2020/06/08 | 2020/12/15 | If the positive number of passengers on the same entry flight reached 5 after entry, the flight would be suspended for 1 week; 10 people were diagnosed, suspended for 4 weeks. | <http://www.caac.gov.cn/XXGK/XXGK/TZTG/202006/t20200604_202928.html> |
|  |  | 2 | 2020/12/16 | 2021/04/30 | If the positive number of passengers on the same entry flight reached 5 after entry, the flight would be suspended for 2 weeks; 10 people were diagnosed, suspended for 4 weeks. | <http://www.caac.gov.cn/XXGK/XXGK/TZTG/202012/t20201216_205607.html> |
|  |  | 3 | 2021/05/01 | Now | If the positive number of passengers on the same entry flight reached 5 after entry, the flight would be suspended for 2 weeks or restrict the flight to 40% load for 4 weeks; 10 people were diagnosed, the flight would be suspended for 4 weeks; 10 people were diagnosed in two consecutive shifts, the flight would be suspended for 8 weeks. | <http://www.caac.gov.cn/XXGK/XXGK/TZTG/202104/t20210429_207386.html> |
| **Local NPIs** | | | | | | |
| 1. **Symptom-based surveillance** | | | | | | |
| Symptom surveillance at fever clinics | Shanghai | - | Before 2022 | Now | Routine monitoring and screening at fever patients | http://shanghai.xinmin.cn/xmsq/2021/08/20/32012714.html |
| Symptom surveillance in the community | Shanghai | - | 2022/2/1 | 2022/2/7 | People from other areas must check health codes and body temperature before entering the community during the Spring Festival | https://baijiahao.baidu.com/s?id=1690818097042939967&wfr=spider&for=pc |
| 2. **Case isolation** | | | | | | |
| Isolation of cases in designated facilities | Shanghai | - | Before 2022 | Now | Several hospitals are set as designated hospitals for medical treatment of COVID-19 cases in Shanghai | https://mp.weixin.qq.com/s/XZLTWktIMLABGZ-9Eum1Qw  https://mp.weixin.qq.com/s/eCQPcwUC9iMirNWmKGGkLw |
| 3. **Contact tracing** | | | | | | |
| Tracing, quarantine and testing of close contacts | Shanghai | - | Before 2022 | Now | 14 days of centralized isolation plus 7 days of home-quarantine, coupled with regular nucleic acid testing | https://mp.weixin.qq.com/s/p1a4A1BrI1q6bZb-IRoxXw |
| Tracing, quarantine and testing of contacts of contacts | Shanghai | - | Before 2022 | Now | 14 days of centralized isolation, coupled with regular nucleic acid testing | https://baijiahao.baidu.com/s?id=1709212142402363456&wfr=spider&for=pc |
| 4. **Occupation-based screening** | Shanghai | - | Before 2022 | Now | Increased frequency of health monitoring and nucleic acid screening will be tailored for the front-line staff at ports, medical staff in fever clinics and other high-risk positions | http://shanghai.xinmin.cn/xmsq/2021/08/20/32012714.html |
| 5. **Targeted screening of individuals at high risk** | Shanghai | - | Before 2022 | Now | Targeted nucleic acid screening for individuals at high risk of exposure, such as close contact, and secondary close contacts, which was identified through field investigation | http://wsjkw.sh.gov.cn/fkdt/20220107/46f73bed4f4d44ea93c4efcdb8008938.html |
| 6. **Social distancing** | | | | | | |
| Travel restrictions for at-risk groups | Shanghai | - | 2020-02-21 | Now | Three-color dynamic management of health code: green code, yellow code and red code. The travel and access to public places will be limited for people with yellow code and red code. | http://sh.bendibao.com/news/2020221/217272.shtm |
|  | Shanghai | - | Before 2022 | Now | All persons coming from or passing through high-risk or medium-risk areas in China should receive centralized quarantine (from high-risk areas) or strict community health monitoring (from medium-risk areas), combining with nucleic acid tests. | https://www.shanghai.gov.cn/sjzccs/20211029/d38089577321468d86b5f0048f419c1f.html |
| Community confinement | At-risk area |  | Before 2022 | Now | Implement "2+12" control measures for the residential community where the close contacts lived or worked. "2" refers to the confinement for 2 days and two nucleic acid tests which have a 24-hour interval at least, and the next stage "12" refers that the community is strictly managed. | https://sghexport.shobserver.com/html/baijiahao/2022/03/14/683716.html  https://new.qq.com/omn/20211203/20211203A05BRO00.html |
| 7. **Other local measures** | | | | | | |
| Universal facemask policies | Shanghai | - | Before 2022 | Now | A graphic version of Guidelines for Wearing Masks in Public Science was released by the National Health Commission of China | https://mp.weixin.qq.com/s/iUPvUHV9GSTEkHMI_xFZiQ |
| **Part II. Additive NPIs of this wave** | | | | | | |
| **Border control measures** | | | | | | |
| Airport diversion of inbound flights | Shanghai | - | 2022/03/21 | 2022/05/01 | The entry point was transferred from Shanghai Pudong Airport to other airports at 12 cities. | http://news.carnoc.com/list/580/580595.html |
| **Local NPIs** | | | | | | |
| 1. **Infection/case isolation** | | | | | | |
| Makeshift isolation hospitals | Shanghai | - | 2022/03/23 | Now | Makeshift isolation hospitals were gradually opened, first adopted at the Minhang District, Shanghai. | https://mp.weixin.qq.com/s/eCQPcwUC9iMirNWmKGGkLw |
| 2. **Mass screening** | | | | | | |
| PCR-based screening of high risk groups | High-risk areas | 1 | 2022/3/16 | 2022/3/17 | 2 nucleic acid screenings within 48 hours, with the adoption of lockdown | https://mp.weixin.qq.com/s/ZJSpoQYjGupSBOsWtSr_vA |
|  | High-risk areas | 2 | 2022/03/23 | 2022/03/24 | 1 nucleic acid screening within 48 hours, with the adoption of lockdown | https://mp.weixin.qq.com/s/VU5d7WNiv7HF6DASXCaYNQ |
|  | High-risk areas | 3 | 2022/03/26 | 2022/3/27 | 1 nucleic acid screening within 48 hours, with the adoption of lockdown | https://mp.weixin.qq.com/s/_OKOPwbPfOw1Zi48FOvF4w |
|  | High-risk areas | 4 | 2022/04/06 | 2022/04/07 | 1 nucleic acid screening in residential communities with positive infections recorded from April 1 to 5, mainly using mixed sampling. | <https://mp.weixin.qq.com/s/QXtV0k0vDYZQkPJ0W2XDDA>; https://mp.weixin.qq.com/s/vU7zkW0SeoqizeU50JLg3g |
|  | Locked-down areas | 5 | 2022/04/18 | 2022/04/21 | 1 nucleic acid test per day | https://mp.weixin.qq.com/s/K-4LVOff1yW9ExUyrwBzlw |
| PCR-based screening of general population | Non-high-risk areas | 1 | 2022/03/18 | 2022/03/20 | 1 nucleic acid screening within 48 hours | https://mp.weixin.qq.com/s/r6OBkpYSHRezqzHXXVclkw |
|  | Eastern Shanghai | 2 | 2022/03/28 | 2022/03/30 | 2 nucleic acid screenings were separately done on March 28 and 30 | https://mp.weixin.qq.com/s/JHHfabFmTO1GQ7hxd5QdYQ |
|  | Western Shanghai | 3 | 2022/04/01 | 2022/04/1 | 1 nucleic acid screening | https://mp.weixin.qq.com/s/jHQoG8YOmCGs6sMxJam_fQ |
|  | Shanghai | 4 | 2022/04/04 | 2022/04/04 | 1 nucleic acid screening | https://mp.weixin.qq.com/s/HWcx2Hv6ONo3tJlduFMYrw |
|  | Shanghai | 5 | 2022/04/10 | 2022/04/10 | 1 nucleic acid screening | https://mp.weixin.qq.com/s/1_3NkXSIKTAUEJBGwIyQkQ |
|  | Shanghai | 6 | 2022/04/26 | 2022/04/26 | 1 nucleic acid screening | https://mp.weixin.qq.com/s/AIlpPep2E1HEC2SEIIT9MA |
|  | Shanghai | 7 | 2022/05/01 | 2022/05/07 | At least 1 nucleic acid screening for the entire Shanghai | https://mp.weixin.qq.com/s/HxQr4RSZIjbwrvoteDM6Vg |
| Antigen-based screening of general population | Non-high-risk areas | 1 | 2022/03/26 | 2022/03/27 | 1 antigen screening | https://mp.weixin.qq.com/s/_OKOPwbPfOw1Zi48FOvF4w |
|  | Shanghai | 2 | 2022/04/03 | 2022/04/03 | 1 antigen screening | https://mp.weixin.qq.com/s/HWcx2Hv6ONo3tJlduFMYrw |
|  | Shanghai | 3 | 2022/04/09 | 2022/04/09 | 1 antigen screening | https://mp.weixin.qq.com/s/L36p7Jrf4HNfARkNzEx-qg |
| 3. **Social distancing** | | | | | | |
| Closure of public places | At-risk area | - | 2022/03/02 | Now | Closure of sports and cultural venues in Putuo District | http://www.xinwenmh.com/38679.html |
| Limiting movement within administrative unit borders | Shanghai | - | 2022/03/12 | Now | No leaving Shanghai unless necessary | https://mp.weixin.qq.com/s/Kev1hnuiRxEqSwjY5J8lpw |
| Limiting gatherings | Shanghai | - | 2022/03/15 | Now | Suspension of gathering activities, such as large-scale exhibitions, cultural performances and others. | https://baijiahao.baidu.com/s?id=1727354297326963152&wfr=spider&for=pc |
| Public transportation closures of long-distance bus | Shanghai | - | 2022/3/14 | Now | The Shanghai Bus Terminals suspend operations | https://mp.weixin.qq.com/s/dku0LLKZUf1hidcQcjCEZQ |
| School closure | Shanghai | 1 | 2022/03/12 | Now | Closure of all kindergartens, nurseries, primary, secondary, vocational, and training schools in Shanghai. | https://mp.weixin.qq.com/s/_yiS-7IhSbp6J9vmQQBYDQ |
|  | Shanghai | 2 | 2022/03/15 | Now | University closure | https://mp.weixin.qq.com/s/_yiS-7IhSbp6J9vmQQBYDQ |
| Grid management of high-risk areas and non-high-risk areas | Shanghai | - | 2022/03/16 | 2022/03/27 | Implemented lockdown for key-areas when launched nucleic acid screening | https://mp.weixin.qq.com/s/ZJSpoQYjGupSBOsWtSr_vA |
| Lockdown | Eastern Shanghai | 1 | 2022/03/28 | 2022/05/31 | Lockdown of eastern Shanghai (which includes the Pudong New District, Fengxian District, Jinshan District, Chongming District, as well as partial Minhang and Songjaing District) | https://mp.weixin.qq.com/s/Ufza89hhBGZsiGPTHoC5aQ |
|  | Western Shanghai | 2 | 2022/04/01 | 2022/05/31 | Lockdown of western Shanghai | https://mp.weixin.qq.com/s/Ufza89hhBGZsiGPTHoC5aQ |

4.	Shanghai Municipal Health Commission. The 134th press conference of COVID-19 in Shanghai. 2022.
